# The Role of Hormonal Contraceptive Use in Mediating Sociodemographic Predictors of Overweight and Obesity among Women of Reproductive Age in Ghana

**DOI:** 10.1101/2025.06.04.25329008

**Authors:** Shaibu Issifu, Lydia Sarponmaa Asante, Issah Sumaila, Debora Awuah Appietuah, Anthony Twum, Helen Agodzo, Michael Opoku-Mireku

## Abstract

**Background:** The global rise in overweight and obesity poses significant public health challenges, contributing to premature mortality and preventable disabilities. Moreover, efforts to increase reproductive autonomy through the promotion of modern contraceptive methods are underway, with persisting concerns regarding their potential influence on weight gain. This study aimed to identify the risk factors associated with overweight and obesity and to examine the mediating role of hormonal contraceptive use among Ghanaian women of reproductive age.

**Methods:** An analytical cross-sectional study was conducted using secondary data from the 2022 Ghana Demographic and Health Survey, which involved 6,181 women aged 20 years and above. Associations between overweight/obesity and various sociodemographic factors were assessed, and logistic regression models were used to estimate crude and adjusted odds ratios. Mediation analysis was performed via structural equation modelling to evaluate the indirect effects of hormonal contraceptive use on the relationships between the independent variables and BMI.

**Results:** The analysis revealed that factors such as age, educational attainment, literacy, marital status, religion, residential status, region, nature of employment, and wealth quintile were significantly associated with higher BMI. The results of the mediation analysis indicated that hormonal contraceptive use partially mediated the relationships between both age (9% of the effect) and marital status (9% of the effect).

**Conclusion:** Hormonal contraceptive use partially mediates the relationship between BMI and both age and marital status, indicating a modest but significant role in weight gain. Integrating individualized weight management counselling into family planning, especially those involving hormonal contraceptives with elevated BMIs, may increase the use of contraceptives while safeguarding women’s health since hormonal contraceptives play an indirect role in weight gain.

## Background

Obesity is characterized by excessive fat accumulation that impairs health [1]. The prevalence of obesity is a significant global health concern because of its association with the pathogenesis of various noncommunicable diseases (NCDs), such as cardiovascular diseases, type 2 diabetes, and certain cancers [2]. Additionally, obesity is the fifth (5^th^) leading risk factor for disease burden globally [3]. The body mass index (BMI), also known as the Quetelet index, is commonly used as a simple and cost-effective measure for classifying overweight and obesity in adults [4],[5]. According to the World Health Organization, in 2021, elevated BMI was responsible for approximately 4 million deaths from NCDs worldwide [6]

The prevalence of obesity has been increasing globally, with more than a billion people classified as obese as of 2022 [6]. This increase is attributed to factors such as the widespread availability of energy-dense [7] foods, nutrient-poor foods, sedentary lifestyles, and urbanization [8]. In particular, women are disproportionately affected [9], partly due to biological factors [10], sociocultural influences [11], inappropriate dietary habits [7] and physical inactivity [12].

Concurrently, efforts to promote modern contraceptive use have been intensified to enhance reproductive health outcomes [13]. Modern contraceptives empower women to make informed decisions about childbearing, thereby preventing unsafe abortion, reducing maternal mortality and improving child health [14]. However, in sub-Saharan Africa, including Ghana, the use of modern contraceptives remains low and is hindered by misconceptions, cultural beliefs, and limited access to healthcare services [15]. Whereas the global uptake is 77% among the target population, sub-Saharan Africa is still lagging, with 56% coverage [13]

One prevalent perception is that hormonal contraceptives lead to significant weight gain, which may deter women from their use [16],[17]. While some studies suggest a potential association between hormonal contraceptive use and weight changes [18], the evidence is inconclusive [19],[20]. Weight changes are often modest [19],[21], with some studies outrightly rejecting a causal association between oral contraceptives and weight gain [22],[23].

Given the increasing obesity rates [6] and the need to improve contraceptive uptake, it is imperative to explore the relationship between BMI and hormonal contraceptive use [16]. Understanding this relationship can inform public health strategies aimed at addressing obesity while promoting effective contraceptive methods. Therefore, this study seeks to examine the determinants of higher BMI and assess the potential mediating role of hormonal contraceptive use among women of reproductive age in Ghana.

## METHODOLOGY

### Study Design

This study analyses secondary data from the nationally representative 2022 Ghana Demographic and Health Survey (GDHS) conducted by the Ghana Statistical Service and partners. The analysis focuses on women aged 20–49 years to examine health and demographic indicators across Ghana’s 16 administrative regions.

### Study Area

Ghana is a West African, English-speaking nation bordered by three French-speaking countries: Burkina Faso to the north, Togo to the east, and Côte d’Ivoire to the west. To the south, it has a coastline along the Gulf of Guinea. Ghana is subdivided into 16 administrative regions. According to the 2021 Population and Housing Census conducted by the Ghana Statistical Service, Ghana has a sex ratio of 97 males for every 100 females and a fertility rate of 3.5% [24].

### Study population

The study population comprised Ghanaian women aged 20 years and above who participated in the 2022 GDHS. This nationally representative survey was conducted by the Ghana Statistical Service (GSS) between October 17, 2022, and January 14, 2023, with technical assistance from the ICF through the DHS Program. While the GDHS primarily targets women aged 15-49 years, this study focuses on women aged 20 years and above and uses the available dataset [25].

### Inclusion and exclusion criteria

1. Women aged 20-49 years

### Study variables

#### Dependent variables

**Overweight/Obesity (High body mass index (BMI)** – A body mass index of 25 kg/m^2^ and above

### Mediating variable

**Hormonal contraceptive usage** – A family planning contraceptive that contains any synthetic hormone (estrogen or progesterone), such as pills, injectables, implants, emergency contraceptive pills, or other modern methods

### Independent variables

#### Sociodemographic characteristics

These variables include age, highest educational level, literacy, residence, region, religion, marital status, wealth quintile, and nature of employment.

#### Sampling Technique

The dataset utilized for this study is the **GHIR8CFL** file from the 2022 GDHS. This dataset comprises information from 15,014 women aged 15-49 years.

To ensure the reliability and generalizability of the findings, the dataset underwent meticulous cleaning via a listwise deletion approach, resulting in a final analytical sample of 6,181 respondents. The cleaning process involved the following steps:

1. Exclusion of Respondents Without Weight or Height Data: A total of 7,338 respondents lacking either weight or height measurements were excluded, as body mass index (BMI) could not be calculated for these individuals.
2. Removal of Respondents Aged Below 20 Years: Given that raw BMI is not an appropriate measure for individuals under 20 years where BMI-for-age is more suitable [26], 1,436 respondents in this age group were excluded.
3. Exclusion of Respondents with Invalid Weights or Height Entries: Fifty-nine (59) respondents with weight or height entries marked as “not present,” “refused,” or “other” were also excluded.

This rigorous cleaning process ensured that the final dataset was robust and suitable for analysing BMI-related outcomes among Ghanaian women aged 20 years and above.

#### Sample size

Following a rigorous data cleaning process, the final analytical sample comprised 6,181 respondents, representing 41.2% of the original dataset. This indicates that 8,833 observations (58.8%) were excluded because of missing or invalid data, as detailed in the data cleaning procedures.

### Data Analysis

The dataset, provided in Stata (.dta) format, was analysed via Stata version 18. Age was categorized into two (2) groups: ideal childbearing age (20-35 years) and above ideal childbearing age (36--49 years), and the 16 administrative regions of Ghana were consolidated into three belts: northern (Northern, Savanna, Upper East, Upper West, and Northeast), middle (Bono, Bono East, Ahafo, Ashanti, and Eastern), and southern (Western, Western North, Central, Greater Accra, Oti, and Volta). Body mass index was computed from weight and height and classified into two categories: normal (<25 kg/m²) and overweight/obese (≥25 kg/m²). Other variables, including religion, were also recategorized into four (Christian, Islamic, traditional, and atheist), and contraceptive use was dichotomized into hormonal and nonhormonal.

The various independent variables used for the study are summarized and presented in a table with their p values determined via the chi-square test with row percentages. The prevalence of overweight/obesity, along with 95% confidence intervals, was estimated via the logit proportion function. Nine (9) independent variables were examined for this study.

Crude odds ratios (cORs) and adjusted odds ratios (aORs) were computed through bivariate and multivariate logistic regression, respectively, for each of the 9 selected predictors to evaluate their strength of association with overweight/obesity. The independent variables were assessed for collinearity via the variable inflation factor (VIF). All the independent variables have VIF values of less than 3 and a mean VIF of 1.54, indicating the absence of collinearity among them.

To investigate the mediating role of hormonal contraceptives in the relationship between BMI and the independent variables, a mediation analysis was conducted. Structural equation modelling (SEM) was employed to construct a directed cyclic graph (DAG) representing the hypothesized mediation pathways. The analysis utilized a nonparametric bootstrap approach with 1,000 resamples and a fixed random seed of 123 to ensure reproducibility. The indirect effects, representing the average causal mediation effects (ACME), were estimated through nonlinear combination analysis. Postestimation bootstrap procedures were applied to obtain bias-corrected coefficients and confidence intervals for the mediation effects. Statistical significance was set at p < 0.05 for all analyses.

### Ethical Issues

Formal authorization was obtained from the Demographic and Health Survey Program for the utilization of the dataset in this study. The GSS obtained clearance from the Ethical Review Committee (ERC) of the Ghana Health Service to ensure that the survey procedures were in accordance with Ghana’s ethical research standards. Also, ICF obtained clearance from the ICF Institutional Review Board (IRB) in accordance with U.S. and international ethical research standards.

## RESULTS

### Sociodemographic characteristics of the respondents

The bivariate analysis in Table 1 below revealed significant associations between all the sociodemographic characteristics and body mass index (BMI) among the study participants (p<0.001). Age was positively associated with high BMI, as approximately 53.5% of individuals above 35 years of age had a high BMI, whereas 38.0% of those aged 20–35 years had a high BMI. Educational attainment was also linked to BMI; 55.7% of respondents with higher education levels had elevated BMIs, which was double the 27.9% observed in individuals without formal education. Literacy levels further influence BMI outcomes. The participants unable to read had the lowest prevalence of high BMI (38.5%). Interestingly, those who could read part of a sentence had a slightly higher prevalence (49.6%) than those who could read an entire sentence (49.0%). Marital status was another significant factor; individuals previously in a union but currently in no relationship had the highest prevalence of high BMI at 55.9%, whereas those never in a union had the lowest at 35.4%.

**Table 1:**
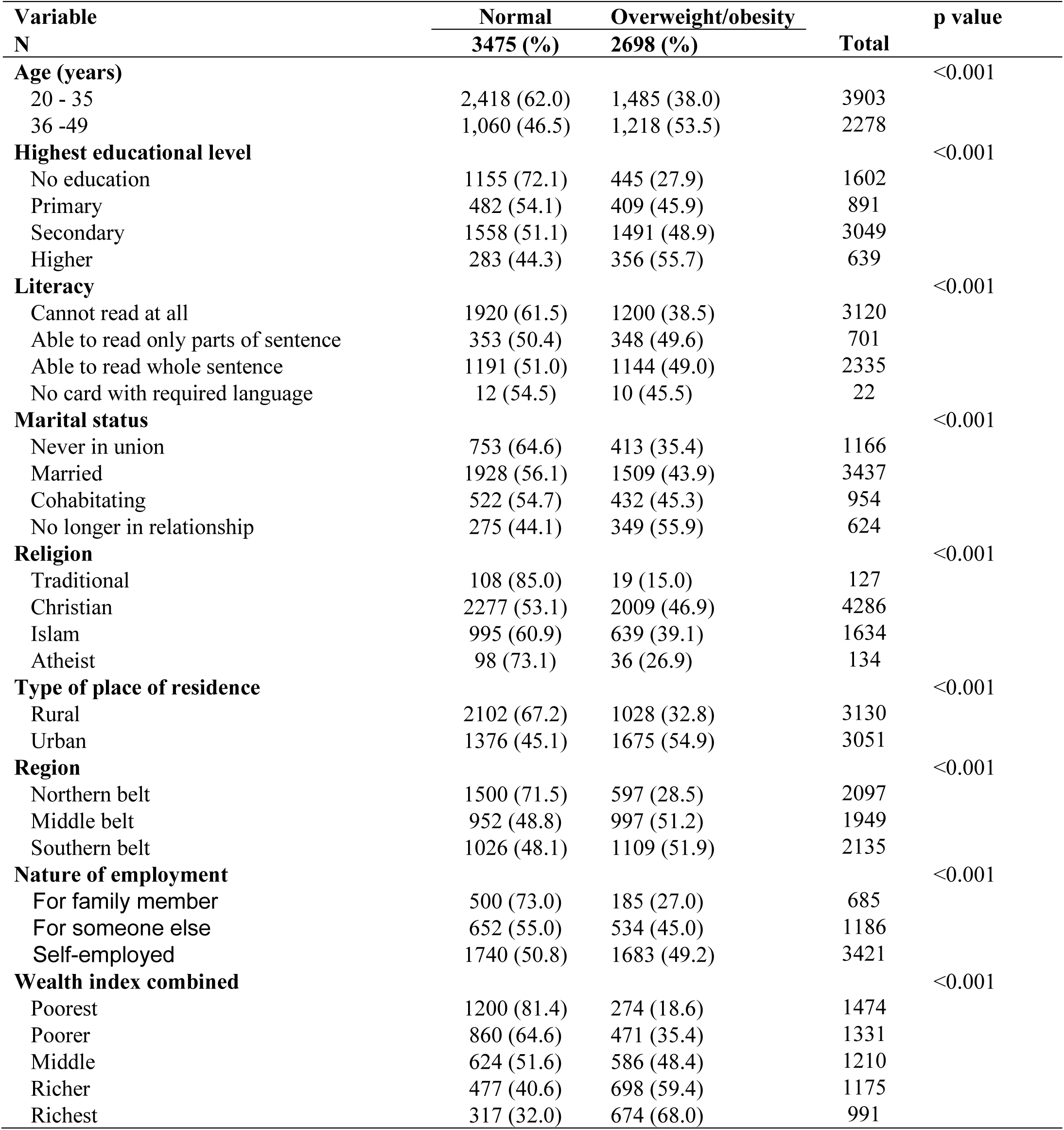
Association between body mass index and the sociodemographic characteristics.

| **Variable** | **Normal** | **Overweight/obesity** |  | **p value** |
| --- | --- | --- | --- | --- |
| **N** | **3475 (%)** | **2698 (%)** | **Total** |  |
| **Age (years)** |  |  |  | <0.001 |
| 20 - 35 | 2,418 (62.0) | 1,485 (38.0) | 3903 |  |
| 36 -49 | 1,060 (46.5) | 1,218 (53.5) | 2278 |  |
| **Highest educational level** |  |  |  | <0.001 |
| No education | 1155 (72.1) | 445 (27.9) | 1602 |  |
| Primary | 482 (54.1) | 409 (45.9) | 891 |  |
| Secondary | 1558 (51.1) | 1491 (48.9) | 3049 |  |
| Higher | 283 (44.3) | 356 (55.7) | 639 |  |
| **Literacy** |  |  |  | <0.001 |
| Cannot read at all | 1920 (61.5) | 1200 (38.5) | 3120 |  |
| Able to read only parts of sentence | 353 (50.4) | 348 (49.6) | 701 |  |
| Able to read whole sentence | 1191 (51.0) | 1144 (49.0) | 2335 |  |
| No card with required language | 12 (54.5) | 10 (45.5) | 22 |  |
| **Marital status** |  |  |  | <0.001 |
| Never in union | 753 (64.6) | 413 (35.4) | 1166 |  |
| Married | 1928 (56.1) | 1509 (43.9) | 3437 |  |
| Cohabiting | 522 (54.7) | 432 (45.3) | 954 |  |
| No longer in relationship | 275 (44.1) | 349 (55.9) | 624 |  |
| **Religion** |  |  |  | <0.001 |
| Traditional | 108 (85.0) | 19 (15.0) | 127 |  |
| Christian | 2277 (53.1) | 2009 (46.9) | 4286 |  |
| Islam | 995 (60.9) | 639 (39.1) | 1634 |  |
| Atheist | 98 (73.1) | 36 (26.9) | 134 |  |
| **Type of place of residence** |  |  |  | <0.001 |
| Rural | 2102 (67.2) | 1028 (32.8) | 3130 |  |
| Urban | 1376 (45.1) | 1675 (54.9) | 3051 |  |
| **Region** |  |  |  | <0.001 |
| Northern belt | 1500 (71.5) | 597 (28.5) | 2097 |  |
| Middle belt | 952 (48.8) | 997 (51.2) | 1949 |  |
| Southern belt | 1026 (48.1) | 1109 (51.9) | 2135 |  |
| **Nature of employment** |  |  |  | <0.001 |
| For family member | 500 (73.0) | 185 (27.0) | 685 |  |
| For someone else | 652 (55.0) | 534 (45.0) | 1186 |  |
| Self-employed | 1740 (50.8) | 1683 (49.2) | 3421 |  |
| **Wealth index combined** |  |  |  | <0.001 |
| Poorest | 1200 (81.4) | 274 (18.6) | 1474 |  |
| Poorer | 860 (64.6) | 471 (35.4) | 1331 |  |
| Middle | 624 (51.6) | 586 (48.4) | 1210 |  |
| Richer | 477 (40.6) | 698 (59.4) | 1175 |  |
| Richest | 317 (32.0) | 674 (68.0) | 991 |  |

Christians had the highest prevalence of high BMI (46.9%), followed by Muslims (39.1%) and traditionalists (15.0%). Compared with rural residency, urban residency was associated with a higher BMI (32.8% vs 54.9%). Regionally, the middle and southern belts reported higher proportions of individuals with elevated BMIs (51.2% and 51.9%, respectively) than did the northern belt (28.5%).

In terms of the nature of employment, self-employed individuals presented a higher prevalence of high BMI, at 49.2%. Additionally, there was a consistent increase in the prevalence of high BMI across ascending wealth quintiles, ranging from 18.6% in the poorest group to 68.0% in the richest group.

### The determinants of body mass index (BMI)

Table 2 below shows the fitting of the crude and adjusted odds ratios to measure the strength of the associations between body mass index and the independent variables. All the variables showed various degrees of association with BMI. Holding all other variables constant, the odds of overweight/obesity were approximately 70% greater (aOR: 1.71, 95% CI: 1.48, 1.95, p < 0.001) for respondents 36–49 years than for those younger than 36 years. After adjusting for confounders, compared with those who had no education, the odds of having a high BMI among those who had various educational levels were as follows: primary education (aOR: 1.70, 95% CI: 1.38, 2.10, p < 0.001); secondary education (aOR: 1.88, 95% CI: 1.52, 2.34, p < 0.001); and higher education (aOR: 2.08, 95% CI: 1.51, 2.88, p < 0.001). Compared with those of respondents who had never been in any union, the odds of having a high BMI were approximately 70% and 57% greater for respondents who were married (aOR: 1.70, 95% CI: 1.405, 2.042, p < 0.001) and cohabitating (aOR: 1.57, 95% CI: 1.257, 1.939, p < 0.001), respectively. The odds were, however, 2 times greater among respondents who were no longer in any relationship (aOR: 2.01; 95% CI: 1.57, 2.58; p < 0.001). In terms of religion, whereas Christians had 2.4 times greater odds of being overweight/obese (aOR: 2.42, 95% CI: 1.36, 4.30; p = 0.003), Muslims had 3.3 times (aOR: 3.28, 95% CI: 1.83, 5.87; p<0.001) greater odds than traditionalists did. Being in an urban residence was associated with a higher BMI, with approximately 23% greater odds than being in a rural residence (aOR: 1.23, 95% CI: 1.06, 1.42; p = 0.005). Additionally, respondents in the middle (aOR: 1.76, 95% CI: 1.47, 2.11, p < 0.001) and southern belts (aOR: 1.57, 95% CI: 1.31, 1.89, p < 0.001) of Ghana had 76% and 57% higher odds of overweight/obesity, respectively, than did respondents in the northern belt. Holding all other factors constant, the odds of overweight/obesity were 53% greater among respondents who were self-employed than among those who were working for family members (aOR: 1.53, 95% CI: 1.25, 1.86, p < 0.001); however, there was no evidence of a significant difference between respondents who worked for other people and those who worked for their family members. When adjusted for confounders, the odds of overweight/obesity increased with increasing wealth compared with the poorest as follows: poorer (aOR: 1.81, 95% CI: 1.48, 2.21, p < 0.001); middle (aOR: 2.72, 95% CI: 2.18, 3.39, p < 0.001), richer (aOR: 4.35, 95% CI: 3.43, 5.53, p < 0.001), and richest (aOR: 6.45, 95% CI: 4.91, 8.49, p < 0.001).

**Table 2:**
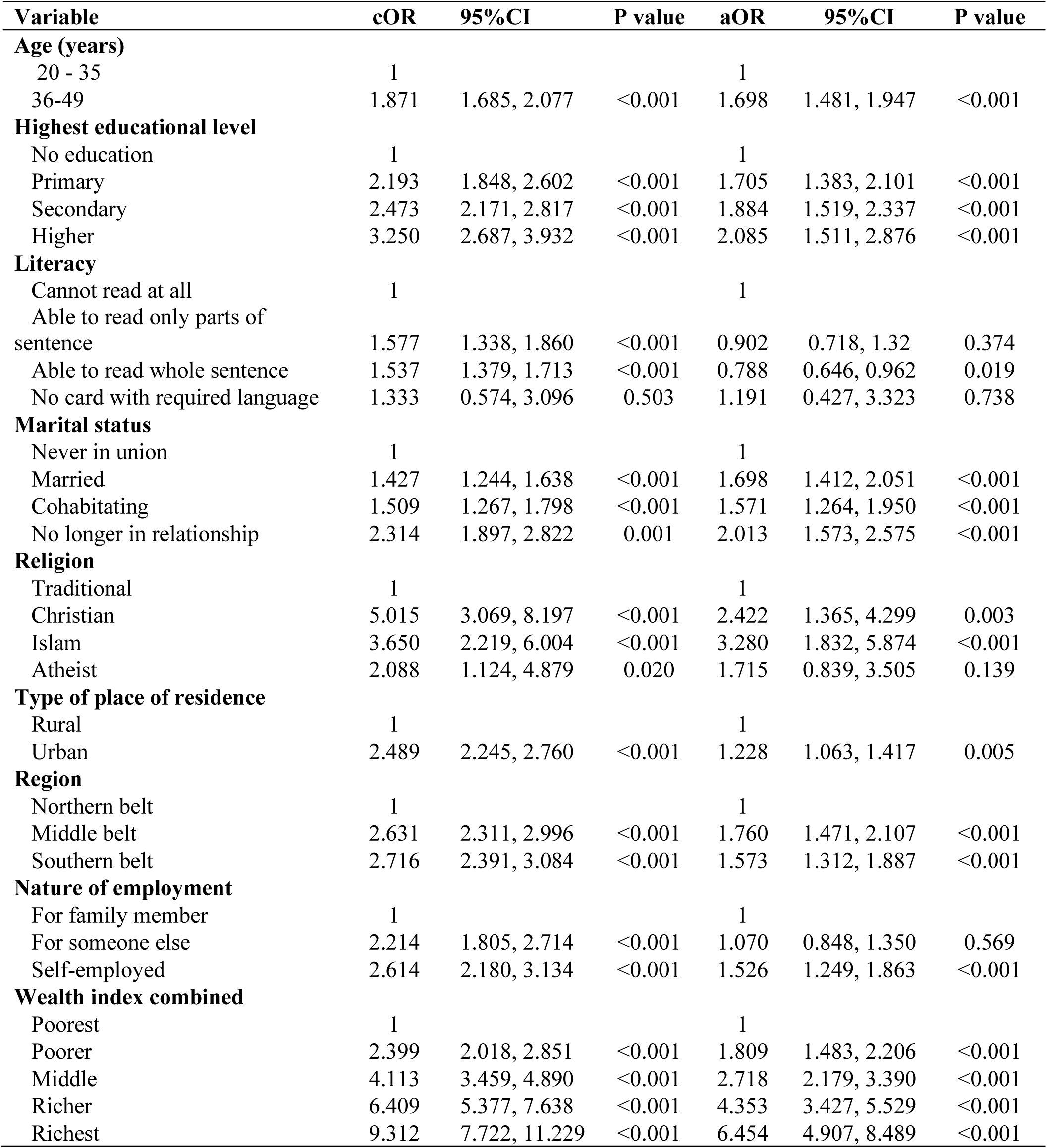
Bivariate and multivariate analyses of body mass index (BMI) and its covariates.

| Variable | cOR | 95%CI | P value | aOR | 95%CI | P value |
| --- | --- | --- | --- | --- | --- | --- |
| **Age (years)** |  |  |  |  |  |  |
| 20 - 35 | 1 |  |  | 1 |  |  |
| 36-49 | 1.871 | 1.685, 2.077 | <0.001 | 1.698 | 1.481, 1.947 | <0.001 |
| **Highest educational level** |  |  |  |  |  |  |
| No education | 1 |  |  | 1 |  |  |
| Primary | 2.193 | 1.848, 2.602 | <0.001 | 1.705 | 1.383, 2.101 | <0.001 |
| Secondary | 2.473 | 2.171, 2.817 | <0.001 | 1.884 | 1.519, 2.337 | <0.001 |
| Higher | 3.250 | 2.687, 3.932 | <0.001 | 2.085 | 1.511, 2.876 | <0.001 |
| **Literacy** |  |  |  |  |  |  |
| Cannot read at all | 1 |  |  | 1 |  |  |
| Able to read only parts of sentence | 1.577 | 1.338, 1.860 | <0.001 | 0.902 | 0.718, 1.32 | 0.374 |
| Able to read whole sentence | 1.537 | 1.379, 1.713 | <0.001 | 0.788 | 0.646, 0.962 | 0.019 |
| No card with required language | 1.333 | 0.574, 3.096 | 0.503 | 1.191 | 0.427, 3.323 | 0.738 |
| **Marital status** |  |  |  |  |  |  |
| Never in union | 1 |  |  | 1 |  |  |
| Married | 1.427 | 1.244, 1.638 | <0.001 | 1.698 | 1.412, 2.051 | <0.001 |
| Cohabiting | 1.509 | 1.267, 1.798 | <0.001 | 1.571 | 1.264, 1.950 | <0.001 |
| No longer in relationship | 2.314 | 1.897, 2.822 | 0.001 | 2.013 | 1.573, 2.575 | <0.001 |
| **Religion** |  |  |  |  |  |  |
| Traditional | 1 |  |  | 1 |  |  |
| Christian | 5.015 | 3.069, 8.197 | <0.001 | 2.422 | 1.365, 4.299 | 0.003 |
| Islam | 3.650 | 2.219, 6.004 | <0.001 | 3.280 | 1.832, 5.874 | <0.001 |
| Atheist | 2.088 | 1.124, 4.879 | 0.020 | 1.715 | 0.839, 3.505 | 0.139 |
| **Type of place of residence** |  |  |  |  |  |  |
| Rural | 1 |  |  | 1 |  |  |
| Urban | 2.489 | 2.245, 2.760 | <0.001 | 1.228 | 1.063, 1.417 | 0.005 |
| **Region** |  |  |  |  |  |  |
| Northern belt | 1 |  |  | 1 |  |  |
| Middle belt | 2.631 | 2.311, 2.996 | <0.001 | 1.760 | 1.471, 2.107 | <0.001 |
| Southern belt | 2.716 | 2.391, 3.084 | <0.001 | 1.573 | 1.312, 1.887 | <0.001 |
| **Nature of employment** |  |  |  |  |  |  |
| For family member | 1 |  |  | 1 |  |  |
| For someone else | 2.214 | 1.805, 2.714 | <0.001 | 1.070 | 0.848, 1.350 | 0.569 |
| Self-employed | 2.614 | 2.180, 3.134 | <0.001 | 1.526 | 1.249, 1.863 | <0.001 |
| **Wealth index combined** |  |  |  |  |  |  |
| Poorest | 1 |  |  | 1 |  |  |
| Poorer | 2.399 | 2.018, 2.851 | <0.001 | 1.809 | 1.483, 2.206 | <0.001 |
| Middle | 4.113 | 3.459, 4.890 | <0.001 | 2.718 | 2.179, 3.390 | <0.001 |
| Richer | 6.409 | 5.377, 7.638 | <0.001 | 4.353 | 3.427, 5.529 | <0.001 |
| Richest | 9.312 | 7.722, 11.229 | <0.001 | 6.454 | 4.907, 8.489 | <0.001 |

### The mediating role of hormonal contraceptives in the relationship between body mass index and its covariates

The mediation analysis shown in Figure 2, depicted in the directed acyclic graph (DAG), explored the relationships between body mass index (BMI) and various sociodemographic factors, with hormonal contraceptive use serving as a potential mediator. A significant direct association was observed between hormonal contraceptive use and BMI (b = 0.040, p = 0.009). Age was positively associated with BMI; individuals aged 36–49 years were more likely to have a higher BMI than those aged 20– 35 years were (c_1_ = 0.12, p < 0.001). However, age was negatively associated with hormonal contraceptive use, indicating that older individuals were less likely to use hormonal contraceptives (a_1_ =-0.099, p < 0.001). Marital status also showed a direct positive association with BMI, with individuals in any form of union being more likely to have a higher BMI than those never in a relationship (c_4_ = 0.04, p < 0.001). Additionally, marital status was positively associated with contraceptive use (a_4_ = 0.022, p < 0.001). Notably, there was a significant direct association between all the independent variables and body mass index. In addition to age and marital status, other sociodemographic variables were significantly associated with hormonal contraceptive use: literacy (a_3_ =-0.019, p = 0.037), religion (a_5_ =-0.032, p = 0.004), and wealth quintile (a_9_ =-0.013, p = 0.027).

**Figure 1:**
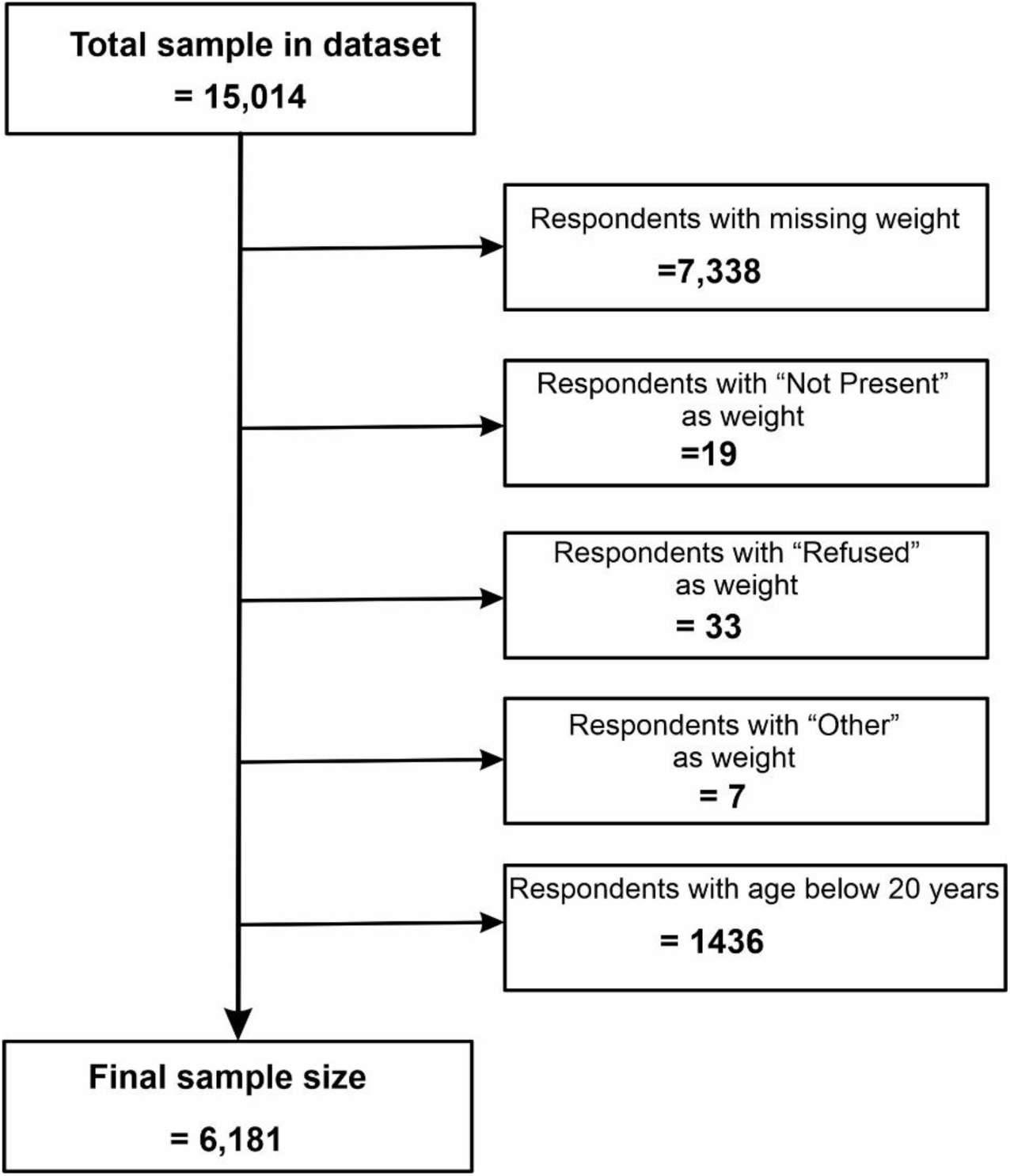
Flow chart of the data cleaning process

**Figure 2:**
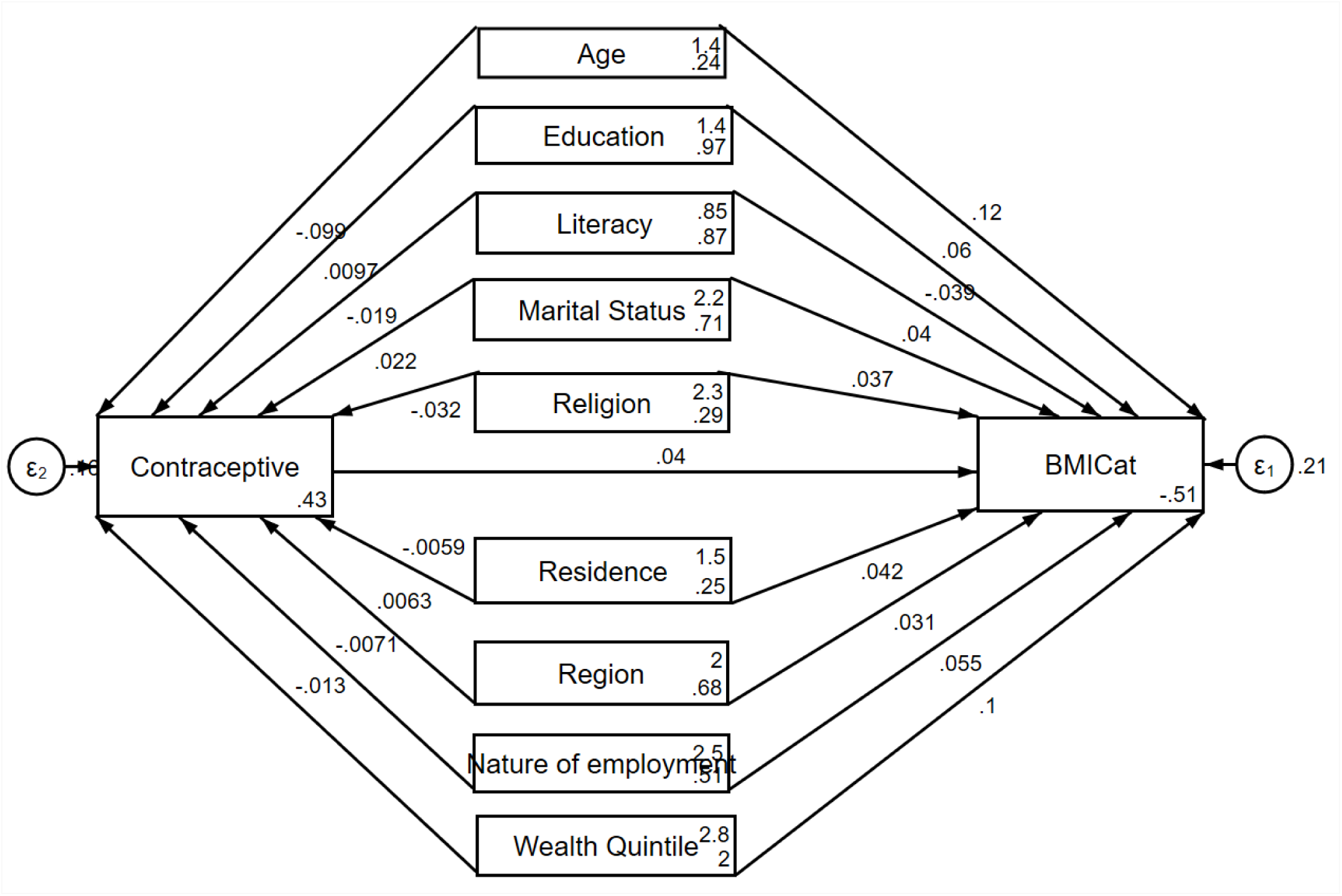
Directed acyclic graph (DAG), showing the indirect relationship between body mass index and its covariate through contraceptives

The results of the mediation analysis presented in Table 3 demonstrate that hormonal contraceptive use significantly mediates the relationships between age, marital status, and body mass index (BMI). Specifically, hormonal contraceptive use partially mediated 9% of the total effect of age on BMI (β₁ =-0.0040, p = 0.012). Additionally, it accounted for 2% of the total effect of marital status on BMI (β₄ = 0.0009, p = 0.042). Other variables, such as education (β_2_ = 0.0004, p = 0.303), literacy (β_3_ = - 0.0008, p = 0.091), religion (β_5_ =-0.0013, p = 0.052), residence (β_6_ =-0.0002, p = 0.669), region (β_7_ = 0.0003, p = 0.429), nature of employment (β_8_ =-0.0003, p = 0.405), and wealth quintile (β_9_ = - 0.0005, p = 0.082), showed nonsignificant mediation effects, with percentages ranging from 0.51% to 3.39%. These findings suggest that hormonal contraceptive use plays a modest but statistically significant mediating role in the associations of age and marital status with BMI.

**Table 3:** Indirect effect of contraceptive use on the relationship between BMI and its factors.

| Variable | Coefficient | P value | [95% conf. Interval] | Percentage mediated |
| --- | --- | --- | --- | --- |
| Age | -0.0040 | **0.012** | -0.0072, -0.0009 | 9.03 |
| Education | 0.0004 | 0.303 | -0.0004, 0.0011 | 0.65 |
| Literacy | -0.0008 | 0.091 | -0.0017, 0.0001 | 1.93 |
| Marital status | 0.0009 | **0.042** | 0.00003, 0.0018 | 2.21 |
| Religion | -0.0013 | 0.052 | -0.0026, 0.00001 | 3.39 |
| Residence | -0.0002 | 0.669 | -0.0013, 0.0008 | 0.55 |
| Region | 0.0003 | 0.429 | -0.0004, 0.0009 | 0.69 |
| Nature of employment | -0.0003 | 0.405 | -0.0010, 0.0004 | 0.52 |
| Wealth Quintile | -0.0005 | 0.082 | -0.0011, 0.0001 | 0.51 |

## DISCUSSION

This study revealed a significant association between higher body mass index (BMI) and various independent variables. To the best of our knowledge, this is the first study to explore the indirect effect of hormonal contraceptive use on the relationship between BMI and the independent variable Ghana. Among the nine independent variables used, hormonal contraceptives partially mediated the relationships between BMI and age or marital status, highlighting their potential role in managing overweight/obesity.

The prevalence of overweight/obesity among the respondents was 43.7% (95% CI: 42.5%, 45.0%), which is consistent with the global prevalence of 43% reported by the WHO [6]. In Ghana, these results confirm the findings of studies by Tette et al. [27] and Lartey et al. [28]. The prevalence was, however, slightly higher than the findings of a study by Obirikorang et al. [29] among undergraduate students and a systematic review and meta-analysis by Yussif et al. [11]. The differences in the results of this study and those of Obirikorang et al. and Yussif et al. could be attributed to the use of a youthful population and the systematic review and meta-analysis, respectively. Compared with their younger counterparts, women aged above 35 years presented 70% greater odds of having an elevated BMI, underscoring the influence of age on BMI. This finding aligns with previous studies conducted in Ghana [30], the United States [31], Pakistan [32], and Spain [33], all of which reported a positive association between increasing age and higher BMI. The observed trend may be attributed to the reduced physical activity associated with aging due to increased responsibilities [34] and economic factors [35].

With respect to literacy, respondents capable of reading a complete sentence had 21% lower odds of having a higher BMI than those unable to read. This finding concurs with the study by Chen et al. [36], which reported a significant negative association between health literacy and obesity in Taiwan.

However, this finding contrasts with the findings of Zare-Zardiny et al. [37], whose study reported a statistically nonsignificant (p = 0.09) association between excess weight and health literacy in Iran. The difference between the studies of Zare-Zardiny and this study may be attributed to geographic disparities. Zare-Zardiny et al. used an adolescent population, whereas this study sampled only women aged 20--49 years. A plausible explanation for the reduced odds of high BMI among the respondents in this study with literacy could be the ability of literates to read, understand and take healthy courses of action, including selecting appropriate foods and engaging in physical activities.

This study revealed a positive association between education and high BMI; the higher the level of education was, the greater the odds of high BMI. The finding of educational status confirms that of John Tetteh et al. in Ghana [27], whose study reported approximately 2 times greater odds among respondents who had completed tertiary education than among those who had not. Additionally, Jimenez-Mora et al. [9] reported an increasing prevalence of overweight with higher education in Colombia. In Iran, Zare-Zardiny et al. [37] reported a positive correlation between educational status and BMI. The likely reason for the higher odds of high BMI for respondents with higher education is their employability, especially those who have completed tertiary education, which equip them financially with the ability to buy what they want [38].

Participants currently or previously in unions were 50% more likely to have a high BMI than those who had never been in a union. This observation is consistent with findings of Nikolic Turnic et al. [39], who reported high BMI among married individuals. Similarly, Amegah et al. [30], in a hospital-based study in Cape Coast, Ghana, noted higher odds of having a high BMI among married respondents. Potential explanations include social support from spouses [35], a sense of security [39], and cultural perceptions valuing larger body sizes within marriages [11].

Urban residents demonstrated increased odds of having an elevated BMI, corroborating global findings [8] and studies from Ghana [27],[40] and Nigeria [41]. The higher odds in urban areas could be a result of the high employment rate in urban areas, which equips them financially to purchase all that they want, including high-caloric foods [38]. This trend may also be linked to increased availability and consumption of ultra-processed foods in urban areas and a high propensity for a sedentary lifestyle [7],[8]. Furthermore, the traditional occupation of rural dwellers is agriculture, which is physically intensive, therefore providing them with the necessary amount of physical activity [8]. To reduce the prevalence of obesity among urban dwellers, there is a need to promote a good built environment that promotes physical activities [42] and even encourages workplace-based physical activities [43]. Additionally, respondents from the middle and southern belts presented 48% and 31% higher odds of having an elevated BMI than did those from the northern belt, respectively. Economic and environmental disparities between these regions may limit access to resources, which tends to be a protective factor for high BMI [44]. For example, the Ghana Standards Living Survey 7 [38] report revealed a very low employment rate in the northern belt (15.5%) compared with the middle belt (26.6%) and southern belt (29.5%), which implies low purchasing power, thereby limiting their ability to purchase and consume highly processed foods.

The wealth quintile also showed a significant association with higher BMI, with odds increasing alongside wealth status, peaking among the richest individuals. This finding aligns with Dogbe [45], who utilized data from the Ghana Living Standards Survey 7. Similar associations have been reported in India [46] and Ethiopia [47]. A plausible explanation is that individuals in higher wealth quintiles have greater purchasing power, potentially leading to increased consumption of calorie-dense foods [48]. This underscores the impact of economic disparities on the prevalence of overweight/obesity.

Importantly, this study revealed the mediating role of hormonal contraceptives in the relationships between BMI and age and marital status. This finding suggests that hormonal contraceptive use plays a modest but statistically significant mediating role in the associations of age and marital status with BMI. This finding aligns with the assertion that the modern family planning method plays a role in weight gain [18]. Whilst the literature on this specific mediation is limited, a study by Avenant et al. established strong associations between hormonal contraceptives and weight gain [20], the study reported a 5% weight increase over baseline after 12 months of etonogestrel implant use, particularly among women under 40 years of age. A longitudinal study reported an average weight gain of 3.8 kg after 24 months of implant use [49]. Rosano et al. [50] highlighted 12 to 24 times greater cardiovascular risks, such as venous thromboembolism, among women with high BMIs on combined oral contraceptives than among those who were not on combined oral contraceptives. Conversely, Procter-Gray et al. [19] and Gallo et al. [22] reported no significant associations between oral contraceptive use and weight gain. Notably, Weidlinger et al. [51] reported that estrogen increased resting energy expenditure by up to 208 kcal, potentially offering a protective effect against overweight and obesity [51]. The present study revealed that the indirect effect of contraceptive use on BMI occurs through age and marital status. This underscores the need to target women within the ideal birth age and Ghanaian women in various forms of relationships (married, cohabiting, and no longer in relationship) with interventions to control their BMI. These findings emphasize the need for further research into the mechanisms by which contraceptives influence weight to inform targeted interventions that promote contraceptive uptake while safeguarding women’s health.

### Limitations

This study is limited by BMI’s inability to assess fat distribution or distinguish fat from muscle. Self-reported data may introduce bias, and the lack of waist and hip measures hinders fat distribution analysis. Lastly, the cross-sectional design prevents causal inferences between variables.

## Conclusion

This study underscores the multifaceted determinants of elevated body mass index (BMI) among Ghanaian women of reproductive age and its interplay with hormonal contraceptive use. The key predisposing factors of overweight/obesity include older age (36-49 years), higher education level, low literacy levels, being married, cohabiting, or previously in a relationship, urban residence, higher wealth quintiles, and regional disparities (middle and southern belts). Importantly, hormonal contraceptive use partially mediates the relationships between BMI and age (9% of the effect) and marital status (2% of the effect), highlighting its potential role in the development of overweight and obesity.

These findings suggest that public health interventions aimed at addressing overweight and obesity in Ghana should consider the complex interplay of sociodemographic factors and hormonal contraceptive use. Tailored strategies such as healthy diets and lifestyle modifications aimed at addressing weight gain in women on hormonal contraceptives may be effective. Such strategies can promote contraceptive use, reduce the risk of obesity, and improve overall health outcomes for Ghanaian women, in line with the broader goal of reducing the burden of noncommunicable diseases.

Future research should explore the underlying mechanisms by which hormonal contraceptives influence weight gain and assess the long-term impacts of these factors on women’s health outcomes.

### Financial disclosure

No funding or grants were secured for this study.

## Data Availability

Data will be made available at a reasonable demand. Also, a copy of the dataset has been made available to the journal

